# Characterization of the Second Wave of the COVID-19 Pandemic in India: A Google Trends Analysis

**DOI:** 10.1101/2021.05.19.21257473

**Authors:** Aayush Visaria, Pooja Polamarasetti, Shivani Reddy, Alizah Ali, Fariha R. Hameed, Joel James, Moizz Akhtar, Sumaiya Islam, Priyanka Raju, Rajat Thawani

## Abstract

**Background:** The second wave of the COVID-19 pandemic has led to considerable morbidity and mortality in India, in part due to lack of healthcare access, low health literacy, and poor disease surveillance. In this retrospective, descriptive ecological study, we utilized Google Trends (GT) to characterize the second COVID-19 wave and its association with official case counts based on search terms related to symptoms, testing, disease complications, medications, preventive behaviors, and healthcare utilization.

**Methods:** GT is a publicly available, online tracking system of Google searches. Searches are presented as relative search volumes (RSV) from 0 (least) to 100 (most number of searches). We performed pre-defined Web searches in India from 2/12/2021 to 5/09/2021. We characterized the peak RSV, RSV doubling rates, and Spearman rank correlation of selected search terms with official case counts. We also used date-adjusted linear regression to estimate the association between highly correlated search terms and official case counts. We then qualitatively classified public search queries into thematic groups to better understand public awareness and needs related to COVID-19.

**Results:** We observed that searches for symptoms (most searched terms in order: fever, cough, headache, fatigue, chest pain), disease states (infection, pneumonia), COVID-19-related medications (remdesivir, ivermectin, azithromycin, Fabiflu, dexamethasone), testing modalities (PCR, CT Scan, D-dimer, C-reactive protein, oxygen saturation), healthcare utilization (oxygen cylinders, hospital, physician), and preventive behaviors (lockdown, mask, pulse oximetry, hand sanitizer, quarantine) all demonstrated increases, in line with increases in official case counts. Symptoms, PCR testing, outpatient medications, and preventive behaviors peaked around April 24th, approximately two weeks prior to the peak RSV in official case counts. Contrarily, healthcare utilization factors, including searches for hospital, physicians, beds, disease states, and inpatient medications did not peak until the first week of May. There were highly significant correlations between ‘Coronavirus Disease 2019’ (r=0.959), ‘fever’ (r=0.935), ‘pulse oximetry’ (r=0.952), ‘oxygen saturation’ (r=0.944), ‘C-reactive protein’ (r=0.955), ‘D-Dimer’ (r=0.945), & ‘Fabiflu’ (r=0.943) and official case counts.

**Conclusion:** GT search terms related to symptoms, testing, and medications are highly correlated with official case counts in India, suggesting need for further studies examining GT’s potential use as a disease surveillance and public informant tool for public health officials.

## Introduction

The COVID-19 pandemic has led to considerable morbidity, mortality, and near healthcare system collapse in India. This has been especially evident during the second wave of the COVID-19 pandemic, which has as of May 19th, 2021, resulted in a cumulative 25.5 million cases and over 283,000 reported deaths [1]. A novel B.1.617.2 Indian variant [2], mass gatherings, political rallies, slow governmental response, and considerable healthcare access issues have put India’s COVID-19 situation to the forefront of international media [3-7]. In addition, many experts, including the World Health Organization and Lancet Commission [3,4], have scrutinized data reporting in India due to its lack of comparability with crematorium accounts, ambiguity surrounding suspected case handling, and paradoxically low case fatality rates compared to other countries [5-7].

Real-world, data-driven approaches to understand public response and conduct disease surveillance are of paramount importance. One such tool, Google Trends^™^ (GT), has been successfully used to describe disease epidemics including Ebola, Influenza, and early COVID-19 responses in India [8]. In addition to nearly universal access to Google and real-time data collection, many individuals search Google before accessing healthcare [9], suggesting that we may be able to better capture disease burden, disease processes, and societal response via the GT tool, especially in countries with inadequate public health infrastructure.

The purpose of our retrospective, descriptive, ecological study of India’s second COVID-19 wave is to utilize GT to describe disease burden, symptomatology, and complications, and their associations with public interest in preventive measures, COVID-19 testing, and vaccination. We also determined the correlation of disease surveillance via GT with government-reported case and death rates. Lastly, we qualitatively describe themes of search queries to help clinicians and public officials better inform and address the general public’s questions.

## Methods

### Data source

We utilized GT, a publicly available, online tracking system of Google searches by search terms, topics, geographic region, and date. Rather than presenting absolute numbers of searches, GT displays the relative search volume (RSV) for pre-specified geographic, time, and search terms. The RSV for a time interval is determined as the search volume in that time interval divided by the largest search volume within the pre-specified criteria. It is then indexed from 0 to 100, where 100 is the reference, largest RSV and 0 is used when there is insufficient data. Further information about GT can be found at https://trends.google.com.

### Search Strategy

As our primary interest was to describe the second wave of India’s COVID-19 pandemic, we conducted Google web searches in India, from February 12, 2021 to May 9th, 2021. The analysis was completed from May 11th to May 18th, 2021.

We initially compiled a list of 141 search terms within the following categories: General COVID-19 terms, Symptoms, Disease Process/Complications, Testing, Healthcare Utilization, Medication Use, Festivals/Holidays, Preventive Behaviors, and Control Searches (see Table 1). Search terms were compiled with the help of physicians, researchers, and patients from India. Based on comparative searches within the GT platform, we narrowed down search terms to the top three to five searches within each category. We also utilized generalized ‘topic’ forms of each search term rather than the exact search term in order to capture variations in related searches such as non-English searches, abbreviations, and cultural and geographic colloquialisms. The final terms used are listed in Table 2.

**Table 1.**
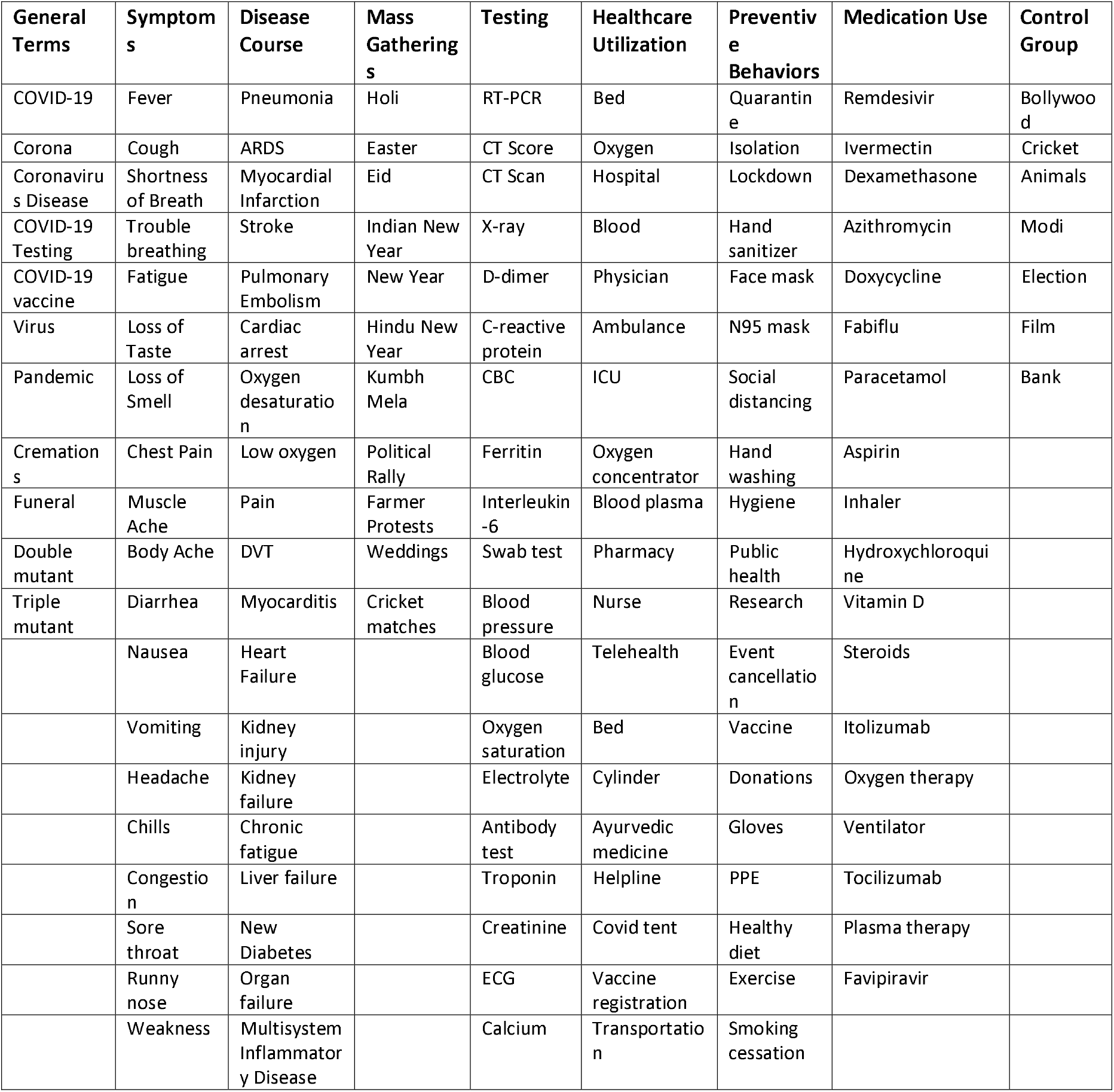
Search Terms by Category

**Table 2.** Top Searched Terms by Category

| **Category** | **Top Searched Topics** |
| --- | --- |
| **General** | Coronavirus Disease 2019, Coronavirus, COVID-19 Testing, COVID-19 Vaccine |
| **Symptoms** | Fever, Cough, Headache, Fatigue, Chest Pain |
| **Disease Process** | Infection, Pneumonia, Myocardial Infarction, Stroke, Pulmonary Embolism |
| **Testing** | Polymerase Chain Reaction, Computed tomography, D-dimer, C-reactive protein, Oxygen Saturation |
| **Healthcare Utilization** | Oxygen Cylinder, Physician, Hospital, Pharmacy, Bed |
| **Medication Use** | Remdesivir, Ivermectin, Azithromycin, Ivermectin, Dexamethasone |
| **Festivals** | Holi, Eid, Indian New Year, Kumbh Mela |
| **Preventive Behaviors** | Lockdown, Quarantine, Mask, Pulse Oximetry, Hand Sanitizer |
| **Control Groups** | Cricket, Bollywood, Animals, Bank, Election |

### Data Management

Once individual searches were completed, the corresponding CSV files were downloaded and combined in order to allow for further data analysis and visualization. This study was exempt from Rutgers IRB approval as all data was publicly available, only at the regional level, and de-identified. In order to compare results with official COVID-19 case and fatality data, we utilized raw data compiled by Johns Hopkins University Center for Systems Science and Engineering (JHU CCSE), available publicly at https://github.com/CSSEGISandData/COVID-19/. Our specific dataset is available upon reasonable request.

### Statistical Analysis

This retrospective, ecological study was primarily meant to be a descriptive analysis of GT data. We determined relative proportional increases in RSV throughout the 90-day time period for all search terms using 2/12/21 as the reference, determined peaks in searches, and described common COVID-19 related questions related to the searches. We also determined initial and second doubling dates: initial or first doubling date was defined as the date during which the three-day average RSV doubled from the average RSV from 2/12/21 to 2/14/21. The second doubling date was the date where the three-day average RSV doubled from the initial doubling date.

We then determined the Spearman rank correlation between official COVID-19 case data and search terms in order to determine whether searches were comparable to official data. Where they were comparable, we subsequently estimated beta coefficients using time-adjusted linear regression to determine significant search covariates. We also performed multivariate linear regression to estimate future caseload; however, due to significant multicollinearity among search terms, this was not statistically sound and thus results are not presented. All analyses were done using SAS 9.4 with a significance level of 0.05.

## Results

### Clinically Relevant Search Terms

Out of 141 search queries, the top topics searched are presented in Table 2. In order from most to least common, searches for symptoms included fever, cough, headache, chest pain, and fatigue (Figure 1a). Whereas the peak RSVs for presenting symptoms (e.g. fever, cough) and associated symptoms (e.g. headache, fatigue) were generally in the last week of April, ‘chest pain’ peaked in the last week of the analysis (Table 3; first week of May). This was in line with official COVID-19 case counts, which also peaked from May 5th - May 7th within the time period of our analysis. Searches for ‘fever’ doubled by March 29th and doubled again 15 days later. This was nearly two weeks after official case counts doubled, whereas the peak RSV for ‘fever’ was two weeks earlier than official counts. The date by which ‘cough’ searches doubled was delayed by approximately 2 weeks compared to ‘fever’ but also demonstrated a second doubling time of 11 days, identical to official case count doubling times (Table 3). Both ‘headache’ and ‘fatigue’ searches did not have a second doubling time due to a high baseline RSV.

**Table 3.** Graphical Characteristics of Top Search Queries

| Search Term | Peak RSV* | Peak 3-day RSV | Initial Doubling Date<sup>#</sup> | 2nd Doubling Date<sup>#</sup> | Case Lag Time<sup>s</sup> |
| --- | --- | --- | --- | --- | --- |
| Official COVID-19 Cases | 5/6/2021 | 5/5 - 5/7 | 3/10/2021 | 3/21/2021 |  |
| **General Terms** |  |  |  |  |  |
| Coronavirus Disease 2019 | 5/1/2021 | 4/29 - 5/1 | 2/28/2021 | 4/2/2021 | 5 |
| Coronavirus | 4/26/2021 | 4/26 - 4/28 | 3/17/2021 | 4/5/2021 | 10 |
| COVID-19 Testing | 4/27/2021 | 4/27 - 4/29 | 3/17/2021 | 3/30/2021 | 9 |
| COVID-19 Vaccine | 5/1/2021 | 4/29 - 5/1 | 2/28/2021 | 4/22/2021 | 5 |
| **Symptoms** |  |  |  |  |  |
| Fever | 4/24/2021 | 4/24 - 4/26 | 3/29/2021 | 4/13/2021 | 12 |
| Headache | 4/26/2021 | 4/29 - 5/1 | 4/23/2021 | N/A<sup>%</sup> | 10 |
| Fatigue | 4/21/2021 | 4/19 - 4/21 | 4/18/2021 | N/A | 15 |
| Cough | 4/28/2021 | 4/21 - 4/23 | 4/10/2021 | 4/21/2021 | 8 |
| Chest pain | 5/5/2021 | 5/5 - 5/7 | 4/8/2021 | 5/2/2021 | 1 |
| **Disease Process/Complications** |  |  |  |  |  |
| Stroke | 5/7/2021 | 3/8 - 3/10 | N/A | N/A | -1 |
| Pneumonia | 5/1/2021 | 5/1 - 5/3 | 4/1/2021 | 4/18/2021 | 5 |
| Myocardial Infarction | 4/30/2021 | 4/29 - 5/1 | 4/28/2021 | N/A | 6 |
| Infection | 5/5/2021 | 5/5 - 5/7 | 4/15/2021 | N/A | 1 |
| **Healthcare Utilization** |  |  |  |  |  |
| Pharmacy | 4/24/2021 | 4/22 - 4/24 | N/A | N/A | 12 |
| Physician | 5/8/2021 | 5/4 - 5/6 | N/A | N/A | -2 |
| Oxygen | 4/24/2021 | 4/24 - 4/26 | 4/6/2021 | 4/13/2021 | 12 |
| Hospital | 5/1/2021 | 5/7 - 5/9 | 4/17/2021 | N/A | 5 |
| Bed | 5/5/2021 | 5/5 - 5/7 | N/A | N/A | 1 |
| **Preventive Behaviors** |  |  |  |  |  |
| Quarantine | 4/19/2021 | 4/19 - 4/21 | 4/3/2021 | N/A | 17 |
| Pulse Oximetry | 4/24/2021 | 4/23 - 4/25 | 3/16/2021 | 3/30/2021 | 12 |
| Mask | 4/26/2021 | 4/26 - 4/28 | 4/14/2021 | N/A | 10 |
| Lockdown | 4/19/2021 | 4/18 - 4/20 | 2/15/2021 | 2/18/2021 | 17 |
| Hand Sanitizer | 4/27/2021 | 5/6 - 5/8 | 4/1/2021 | 4/14/2021 | 9 |
| **Testing** |  |  |  |  |  |
| Oxygen Saturation | 5/5/2021 | 5/4 - 5/6 | 3/20/2021 | 3/30/2021 | 1 |
| Computed Tomography | 5/3/2021 | 5/2 - 5/4 | 3/29/2021 | 4/14/2021 | 3 |
| Polymerase Chain Reaction | 4/19/2021 | 4/22 - 4/24 | 2/23/2021 | 3/24/2021 | 17 |
| C-reactive Protein | 5/8/2021 | 5/6 - 5/8 | 3/18/2021 | 4/2/2021 | -2 |
| D-Dimer | 5/6/2021 | 5/6 - 5/8 | 3/11/2021 | 3/29/2021 | 0 |
| **Medication Use** |  |  |  |  |  |
| Dexamethasone | 5/1/2021 | 5/2 - 5/3 | 4/8/2021 | 4/18/2021 | 5 |
| Azithromycin | 5/3/2021 | 5/3 - 5/5 | 3/25/2021 | 4/7/2021 | 3 |
| Ivermectin | 4/25/2021 | 4/23 - 4/25 | 3/20/2021 | 4/3/2021 | 11 |
| Fabiflu | 4/23/2021 | 4/23 - 4/25 | 3/15/2021 | 3/22/2021 | 13 |
| Remdesivir | 4/18/2021 | 4/18 - 4/20 | 3/20/2021 | 3/31/2021 | 18 |

Format of dates: Month/Day/Year; RSV = Relative Search Volume. All searches conducted from 2/12/21 to 5/9/21
\*Peak RSV refers to the date at which a specific search query hits 100 (its highest point in our time period)

**Figure 1.**
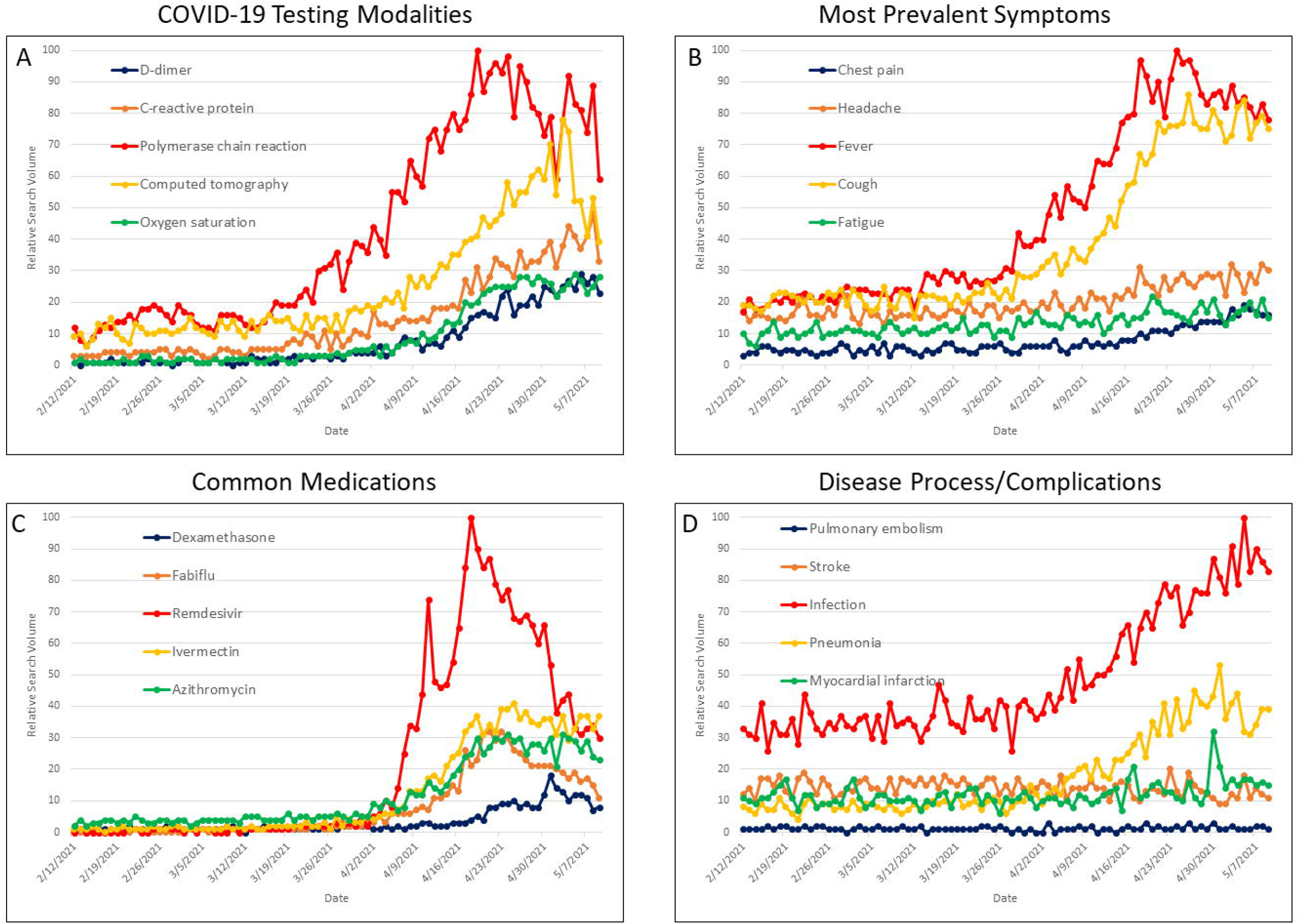
Clinically Relevant COVID-19 Search Terms.

Among disease complications, most individuals searched for pneumonia, infection, myocardial infarction, and stroke, in that order (Figure 1b). Searches for ‘pneumonia’ peaked on May 1st and had a first doubling date of April 1st and a second doubling date of April 18th. This is approximately 4 weeks after official case doubling times. However, the peak RSV for ‘pneumonia’ was five days prior to the peak in official cases (Table 3).

Among testing modalities, searches for ‘polymerase chain reaction’ (PCR) were of the highest RSV, followed by ‘computed tomography’ (CT Scan), ‘C-reactive Protein’, ‘D-dimer’, and ‘Oxygen Saturation’ (Figure 1c). The peak RSV was two weeks earlier for PCR searches than other testing modalities. Compared to official case counts, PCR searches peaked two weeks earlier and also had a first doubling date two weeks earlier (Table 3).

Among medication searches, the majority of searches were for ‘remdesivir’, followed by ‘ivermectin’, ‘azithromycin’, ‘fabiflu’, and ‘dexamethasone’ (Figure 1d). Ivermectin and Fabiflu, both medications used in the outpatient setting, peaked around April 23-25th, with an initial doubling date from March 15-20th. Largely inpatient medications, such as ‘remdesivir’ and ‘dexamethasone’ were more variable in their peak RSV and doubling times, with ‘remdesivir’ parameters two weeks earlier than ‘dexamethasone’ parameters (Table 3).

### Socially-relevant Search Terms

General, healthcare utilization, preventive behavior, and festival/holiday searches also demonstrated similar peak RSVs and doubling times as clinically-relevant search terms (Table 3). General searches for the ‘Coronavirus Disease 2019’ topic peaked on May 1st, had a first doubling date of February 28th, approximately two weeks prior to official case count doubling, and a second doubling date of April 2nd, approximately two weeks after official case count doubling.

Among healthcare utilization searches, searches related to ‘oxygen’ were most prevalent, peaking on April 24th and having a second doubling time within one week of first doubling date. Searches for ‘physician’, ‘hospital’, and ‘bed’ peaked in the first week of May and did not show any doubling (Figure 2b, Table 3).

**Figure 2.**
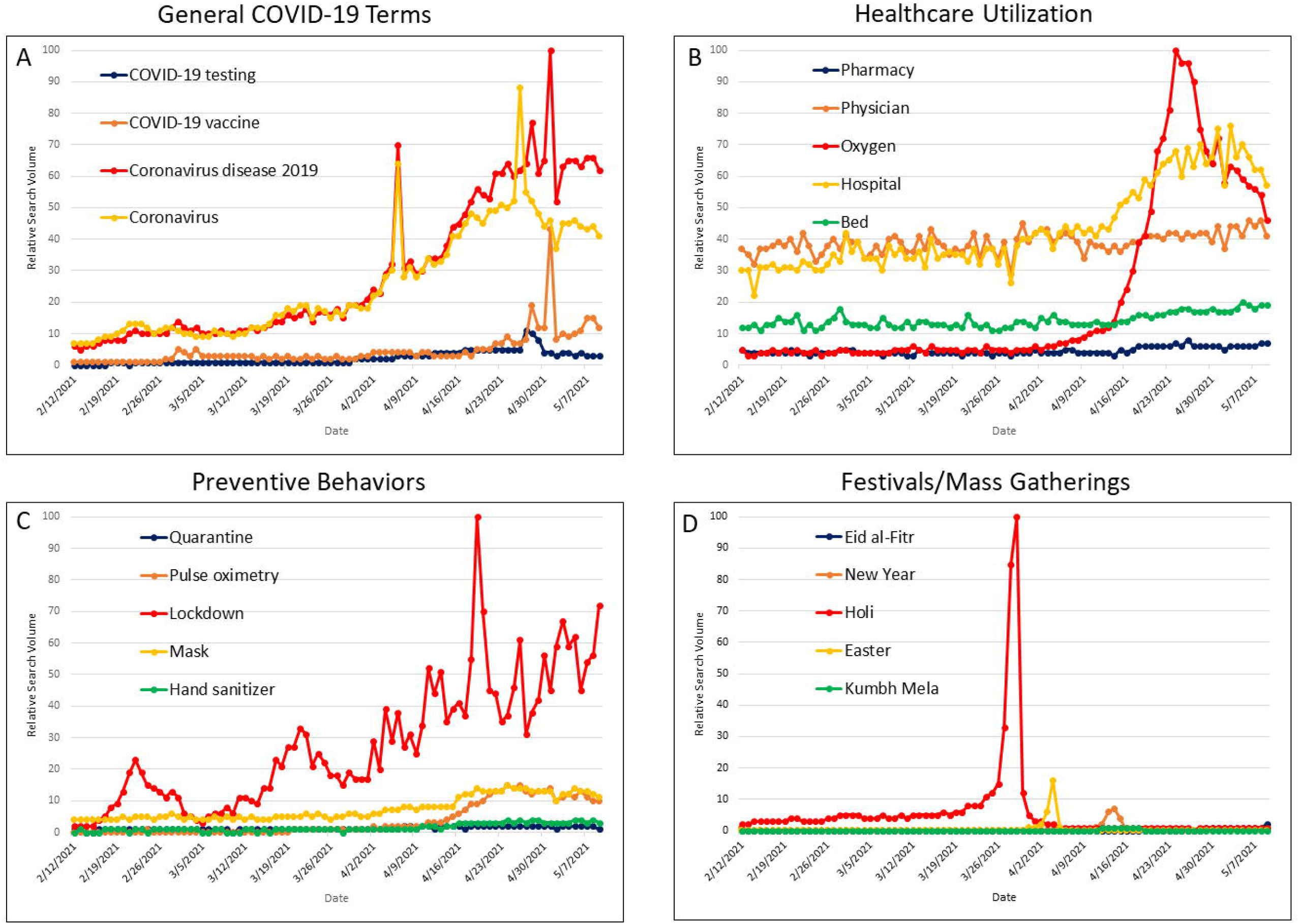
Socially Relevant COVID-19 Search Terms.

Among preventive behavior searches, we observed a general increase in searches for ‘lockdown’, as well as ‘mask’ and ‘pulse oximetry’ (Figure 1c). Nearly all preventive behavior searches peaked from April 18th to April 28th but had variable doubling dates (Table 3).

Among searches for festivals/mass gatherings, we see a tremendous increase in searches for Holi on March 29th and smaller, but significant, increases in other gatherings (Figure 2d).

Peak searches of Eid-al-Fitr, Holi, Easter and Indian New Year occurred in the days leading up to the holiday. No searches under these general categories included queries regarding COVID-19. However, when searching for queries associated with Kumbh Mela, a Hindu festival that was held in Haridwar, Uttarakhand during April 2021 known to have been a “super-spreader” event, had increasing queries regarding the festival and “covid” or “covid cases” during this time frame. When stratifying Indian New Year by regional festivities such as Ugadi, Gudi Padwa, Vaisakhi, and Vishu, there were no searches associated with “covid”.

### Correlation Between Searches and Official Case Counts

We determined the Spearman rank correlation between search terms and official case counts (Table 4). The searches yielding the highest correlation included ‘Coronavirus Disease 2019’ (r=0.959), ‘fever’ (r=0.935), ‘pulse oximetry’ (r=0.952), ‘oxygen saturation’ (r=0.944), ‘C-reactive protein’ (r=0.955), ‘D-Dimer’ (r=0.945), and ‘fabiflu’ (r=0.943). This can also be seen visually in Figure 3. Using date-adjusted linear regression to estimate the association between search terms and official case counts, we found that every 1 unit increase in the RSV for ‘fever’ resulted in a mean increase of 4,854 cases (95% CI: 4,538 - 5,170; p<0.0001). Every 1 unit increase in ‘pulse oximetry’ yielded a mean increase of 3,141 (95% CI: 2,863 - 3,420; p<0.0001) cases. Every 1 unit increase in ‘fabiflu’ resulted in a mean increase of 2,318 (95% CI: 1,821 - 2,815; p<0.0001) cases. When looking at the correlation between official case counts and control search terms, we found that ‘film’ and ‘election’ had no correlation, but other control terms including ‘cricket’, ‘bank’, and ‘animal’ were significantly negatively correlated (Table 4). When we determined the correlation of search terms with official case counts stratified by time period (first 21 days, middle period, and last 21 days), we observed that correlations were of lesser significance in the first and last 21 days (data not shown).

**Table 4.**
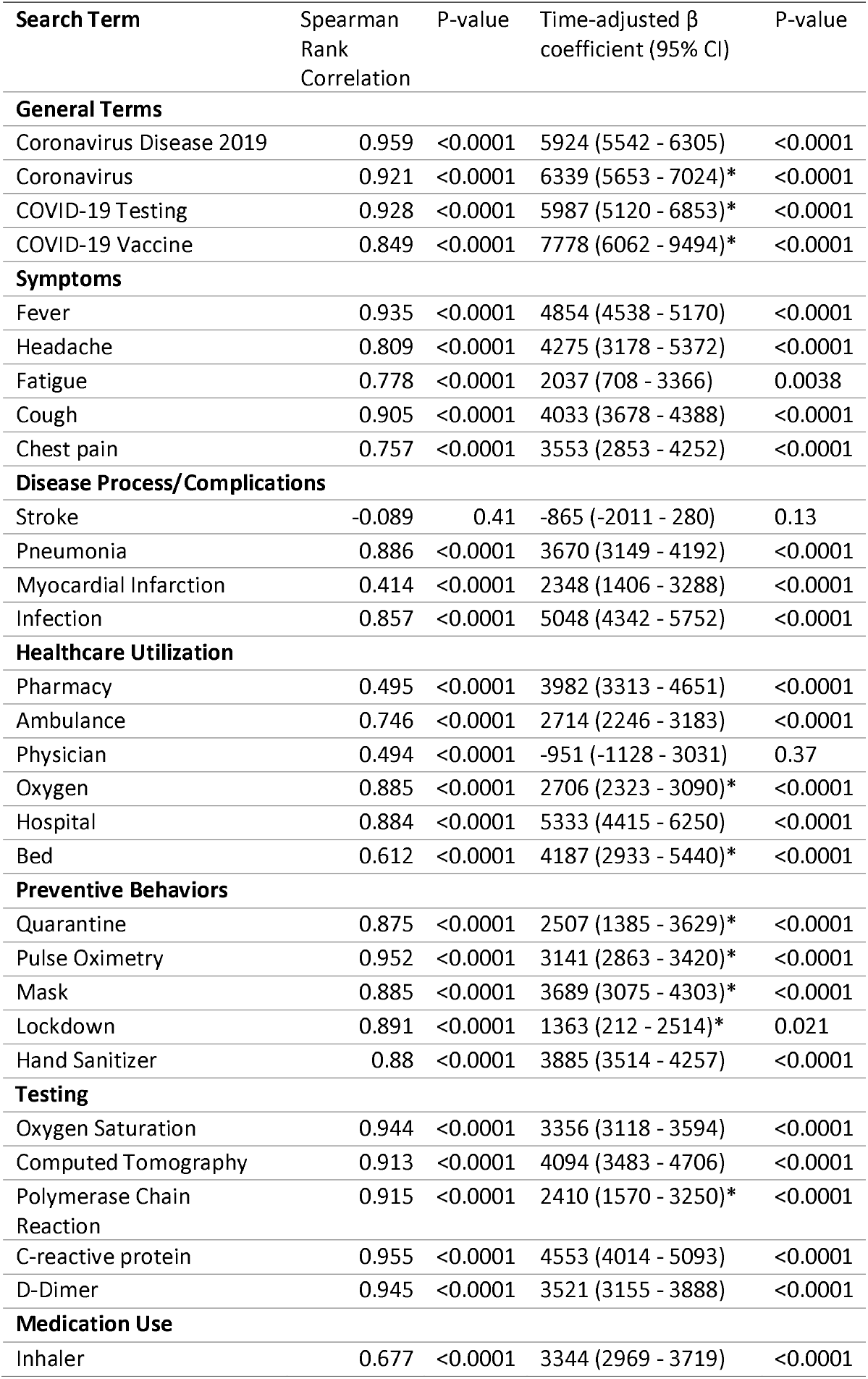
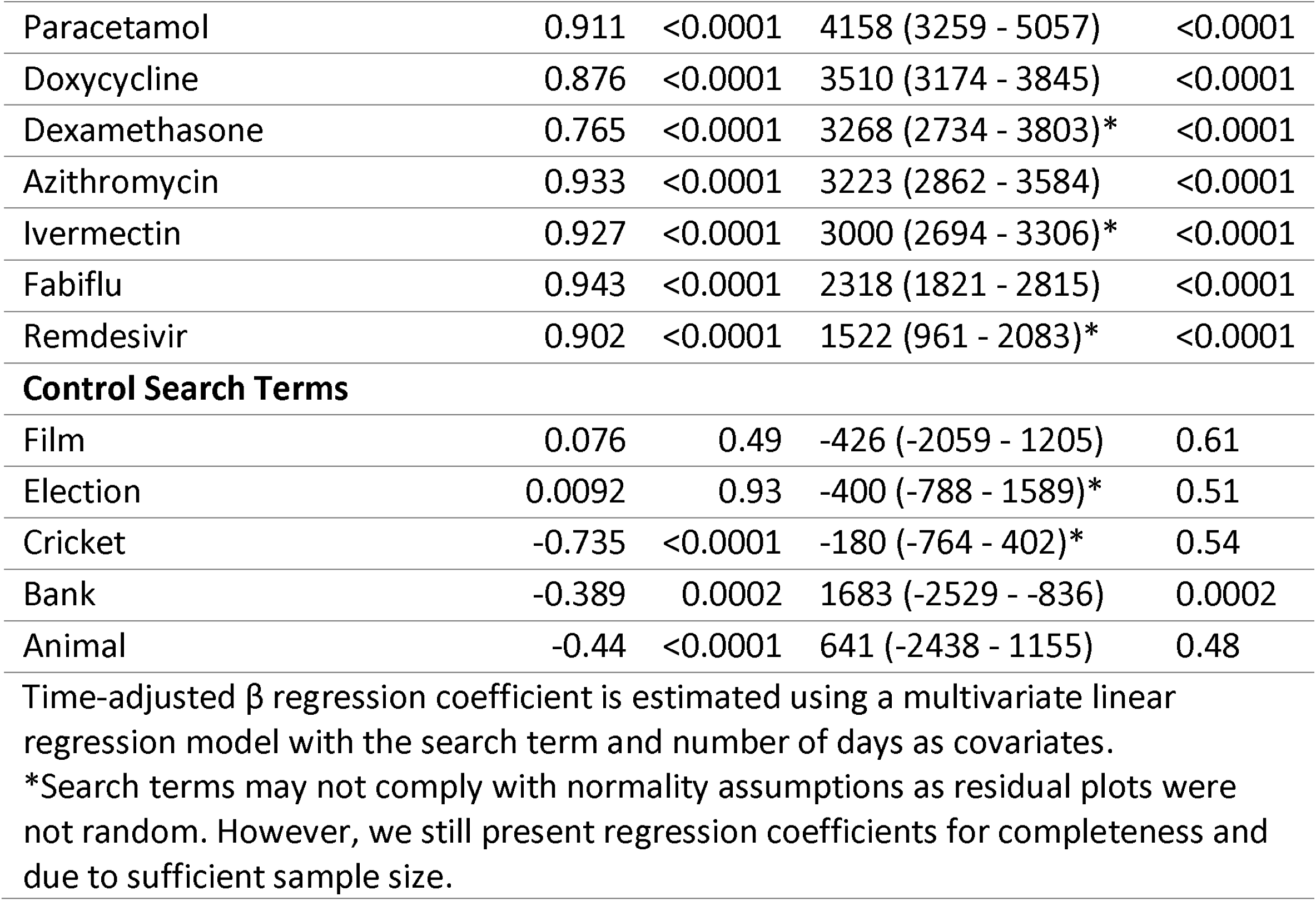
Correlation Between Search Terms and Official Case Count

| Search Term | Spearman Rank Correlation | P-value | Time-adjusted $\beta$ coefficient (95% CI) | P-value |
| --- | --- | --- | --- | --- |
| **General Terms** |  |  |  |  |
| Coronavirus Disease 2019 | 0.959 | <0.0001 | 5924 (5542 - 6305) | <0.0001 |
| Coronavirus | 0.921 | <0.0001 | 6339 (5653 - 7024)* | <0.0001 |
| COVID-19 Testing | 0.928 | <0.0001 | 5987 (5120 - 6853)* | <0.0001 |
| COVID-19 Vaccine | 0.849 | <0.0001 | 7778 (6062 - 9494)* | <0.0001 |
| **Symptoms** |  |  |  |  |
| Fever | 0.935 | <0.0001 | 4854 (4538 - 5170) | <0.0001 |
| Headache | 0.809 | <0.0001 | 4275 (3178 - 5372) | <0.0001 |
| Fatigue | 0.778 | <0.0001 | 2037 (708 - 3366) | 0.0038 |
| Cough | 0.905 | <0.0001 | 4033 (3678 - 4388) | <0.0001 |
| Chest pain | 0.757 | <0.0001 | 3553 (2853 - 4252) | <0.0001 |
| **Disease Process/Complications** |  |  |  |  |
| Stroke | -0.089 | 0.41 | -865 (-2011 - 280) | 0.13 |
| Pneumonia | 0.886 | <0.0001 | 3670 (3149 - 4192) | <0.0001 |
| Myocardial Infarction | 0.414 | <0.0001 | 2348 (1406 - 3288) | <0.0001 |
| Infection | 0.857 | <0.0001 | 5048 (4342 - 5752) | <0.0001 |
| **Healthcare Utilization** |  |  |  |  |
| Pharmacy | 0.495 | <0.0001 | 3982 (3313 - 4651) | <0.0001 |
| Ambulance | 0.746 | <0.0001 | 2714 (2246 - 3183) | <0.0001 |
| Physician | 0.494 | <0.0001 | -951 (-1128 - 3031) | 0.37 |
| Oxygen | 0.885 | <0.0001 | 2706 (2323 - 3090)* | <0.0001 |
| Hospital | 0.884 | <0.0001 | 5333 (4415 - 6250) | <0.0001 |
| Bed | 0.612 | <0.0001 | 4187 (2933 - 5440)* | <0.0001 |
| **Preventive Behaviors** |  |  |  |  |
| Quarantine | 0.875 | <0.0001 | 2507 (1385 - 3629)* | <0.0001 |
| Pulse Oximetry | 0.952 | <0.0001 | 3141 (2863 - 3420)* | <0.0001 |
| Mask | 0.885 | <0.0001 | 3689 (3075 - 4303)* | <0.0001 |
| Lockdown | 0.891 | <0.0001 | 1363 (212 - 2514)* | 0.021 |
| Hand Sanitizer | 0.88 | <0.0001 | 3885 (3514 - 4257) | <0.0001 |
| **Testing** |  |  |  |  |
| Oxygen Saturation | 0.944 | <0.0001 | 3356 (3118 - 3594) | <0.0001 |
| Computed Tomography | 0.913 | <0.0001 | 4094 (3483 - 4706) | <0.0001 |
| Polymerase Chain Reaction | 0.915 | <0.0001 | 2410 (1570 - 3250)* | <0.0001 |
| C-reactive protein | 0.955 | <0.0001 | 4553 (4014 - 5093) | <0.0001 |
| D-Dimer | 0.945 | <0.0001 | 3521 (3155 - 3888) | <0.0001 |
| **Medication Use** |  |  |  |  |
| Inhaler | 0.677 | <0.0001 | 3344 (2969 - 3719) | <0.0001 |
| Paracetamol | 0.911 | <0.0001 | 4158 (3259 - 5057) | <0.0001 |
| Doxycycline | 0.876 | <0.0001 | 3510 (3174 - 3845) | <0.0001 |
| Dexamethasone | 0.765 | <0.0001 | 3268 (2734 - 3803)* | <0.0001 |
| Azithromycin | 0.933 | <0.0001 | 3223 (2862 - 3584) | <0.0001 |
| Ivermectin | 0.927 | <0.0001 | 3000 (2694 - 3306)* | <0.0001 |
| Fabiflu | 0.943 | <0.0001 | 2318 (1821 - 2815) | <0.0001 |
| Remdesivir | 0.902 | <0.0001 | 1522 (961 - 2083)* | <0.0001 |
| **Control Search Terms** |  |  |  |  |
| Film | 0.076 | 0.49 | -426 (-2059 - 1205) | 0.61 |
| Election | 0.0092 | 0.93 | -400 (-788 - 1589)* | 0.51 |
| Cricket | -0.735 | <0.0001 | -180 (-764 - 402)* | 0.54 |
| Bank | -0.389 | 0.0002 | 1683 (-2529 - -836) | 0.0002 |
| Animal | -0.44 | <0.0001 | 641 (-2438 - 1155) | 0.48 |

Time-adjusted $\beta$ regression coefficient is estimated using a multivariate linear regression model with the search term and number of days as covariates.
\*Search terms may not comply with normality assumptions as residual plots were not random. However, we still present regression coefficients for completeness and due to sufficient sample size.

**Figure 3.**
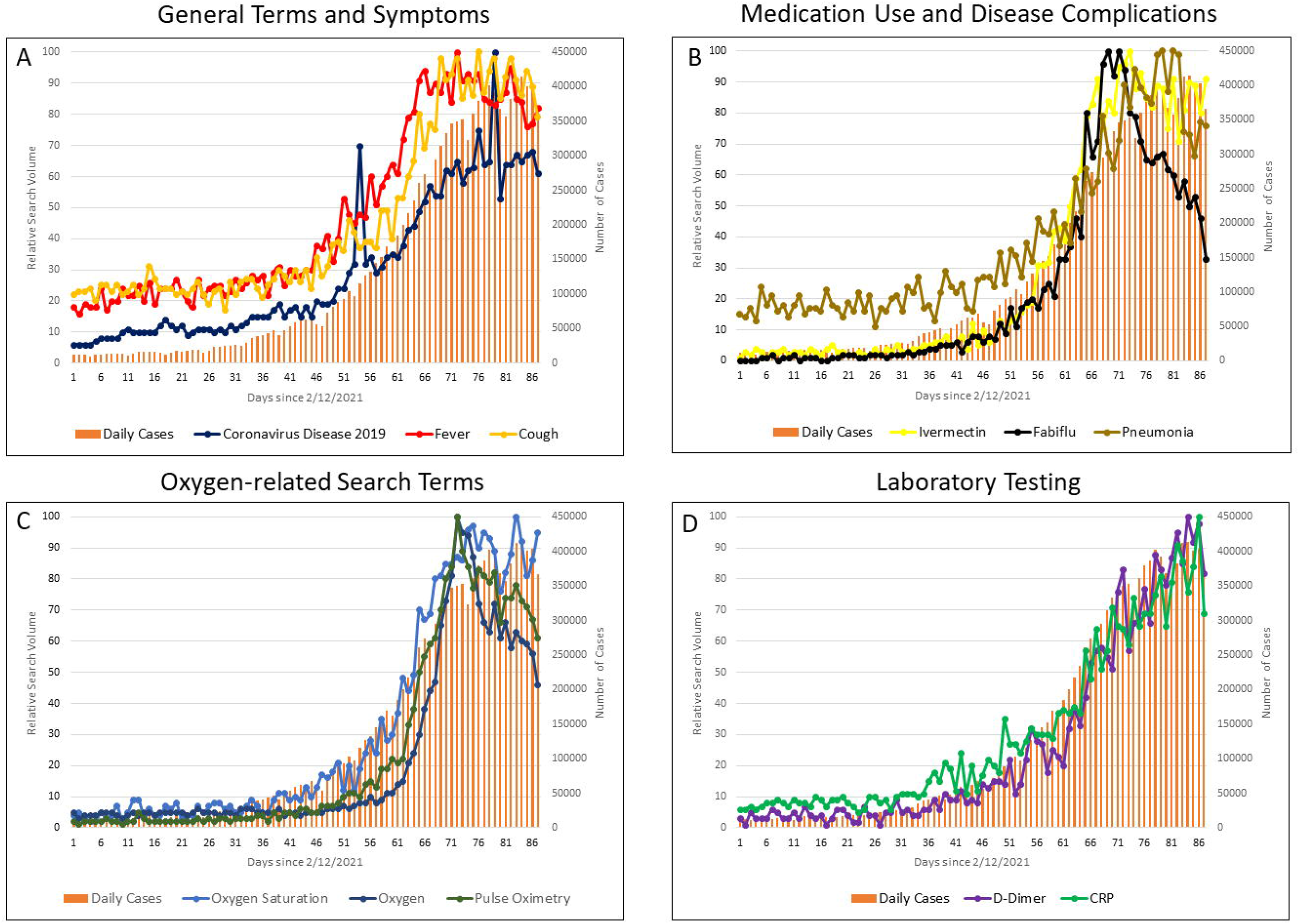
Correlation Between COVID-19 Cases and Search Terms.

We also determined the correlation between search terms and official death counts, which yielded similar but lower magnitude correlations (Table 5).

**Table 5.** Correlation Between Search Terms and Official COVID-19 Death Counts

| Search Term | Spearman Rank Correlation | P-value |
| --- | --- | --- |
| **General Terms** |  |  |
| Coronavirus Disease 2019 | 0.945 | <0.0001 |
| Coronavirus | 0.918 | <0.0001 |
| COVID-19 Testing | 0.916 | <0.0001 |
| COVID-19 Vaccine | 0.816 | <0.0001 |
| **Symptoms** |  |  |
| Fever | 0.921 | <0.0001 |
| Headache | 0.808 | <0.0001 |
| Fatigue | 0.764 | <0.0001 |
| Cough | 0.908 | <0.0001 |
| Chest pain | 0.75 | <0.0001 |
| **Disease Process/Complications** |  |  |
| Stroke | -0.131 | 0.23 |
| Pneumonia | 0.89 | <0.0001 |
| Myocardial Infarction | 0.417 | <0.0001 |
| Infection | 0.853 | <0.0001 |
| **Healthcare Utilization** |  |  |
| Pharmacy | 0.513 | <0.0001 |
| Ambulance | 0.741 | <0.0001 |
| Physician | 0.508 | <0.0001 |
| Oxygen | 0.878 | <0.0001 |
| Hospital | 0.863 | <0.0001 |
| Bed | 0.585 | <0.0001 |
| **Preventive Behaviors** |  |  |
| Quarantine | 0.869 | <0.0001 |
| Pulse Oximetry | 0.954 | <0.0001 |
| Mask | 0.883 | <0.0001 |
| Lockdown | 0.897 | <0.0001 |
| Hand Sanitizer | 0.897 | <0.0001 |
| **Testing** |  |  |
| Oxygen Saturation | 0.941 | <0.0001 |
| Computed Tomography | 0.908 | <0.0001 |
| Polymerase Chain Reaction | 0.918 | <0.0001 |
| C-Reactive Protein | 0.952 | <0.0001 |
| D-Dimer | 0.938 | <0.0001 |
| **Medication Use** |  |  |
| Inhaler | 0.709 | <0.0001 |
| Paracetamol | 0.89 | <0.0001 |
| Doxycycline | 0.864 | <0.0001 |
| Dexamethasone | 0.785 | <0.0001 |
| Azithromycin | 0.929 | <0.0001 |
| Ivermectin | 0.927 | <0.0001 |
| Fabiflu | 0.927 | <0.0001 |
| Remdesivir | 0.891 | <0.0001 |
| **Control Search Terms** |  |  |
| Film | 0.072 | 0.51 |
| Election | -0.012 | 0.91 |
| Cricket | -0.729 | <0.0001 |
| Bank | -0.37 | 0.0004 |
| Animal | -0.448 | <0.0001 |

### Qualitative Theme Analysis

In order to understand the underlying questions for specific search topics, we grouped related search queries into observed themes, namely administrative (testing, registration, location, costs), symptom expectations (disease progression, symptom management), and health literacy (meaning of terms, etiology, normal levels). Many individuals seemingly are unsure of testing locations, vaccine registration protocol, location of available hospitals, and costs of various tests, medications and healthcare utilization factors. Individuals are also unsure of whether presenting symptoms are related to COVID-19 and have questions about expected length of fever, definition of fever, normal values for common laboratory tests, and over-the-counter treatment options for common symptoms like fever and cough (Table 6).

**Table 6:**
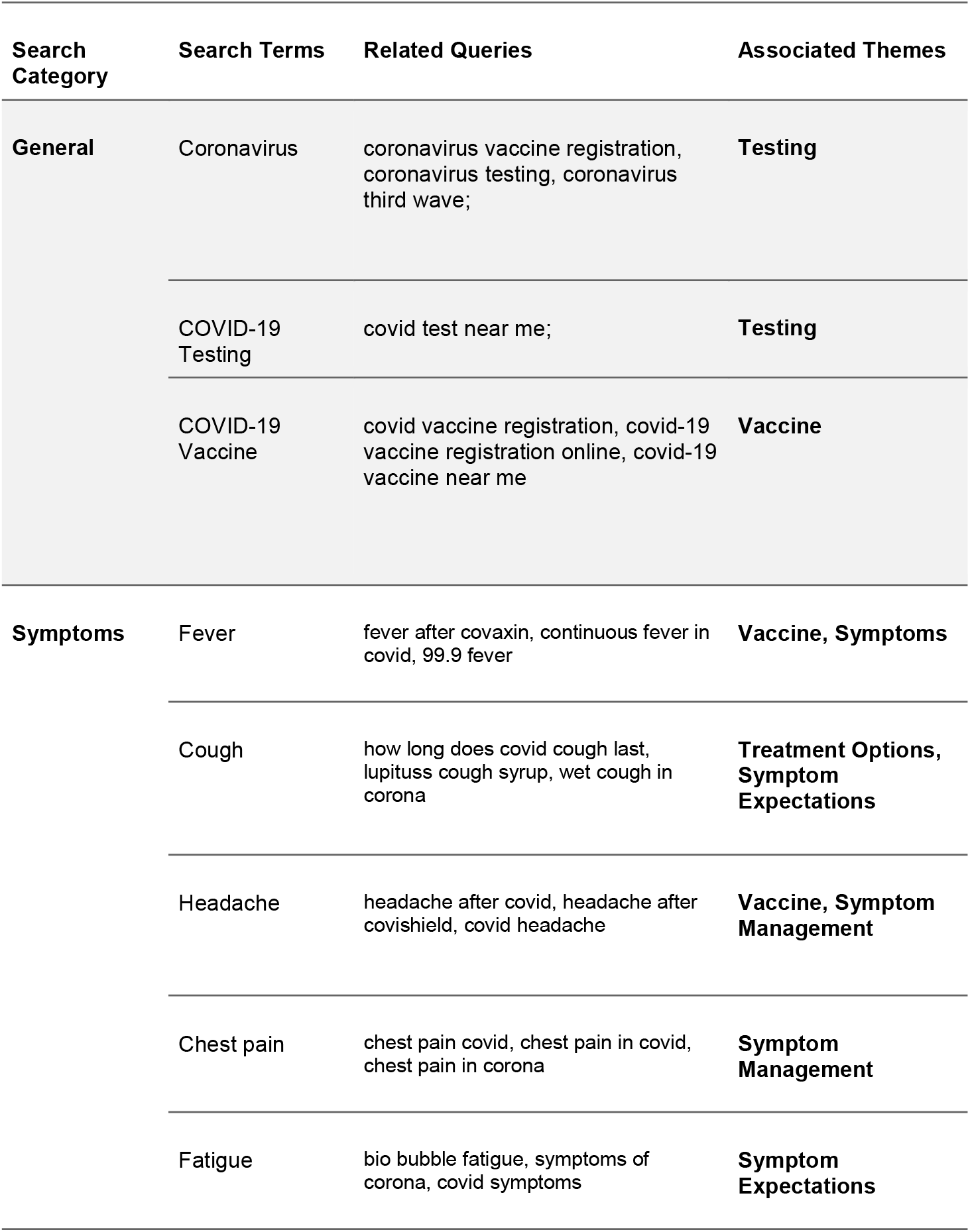
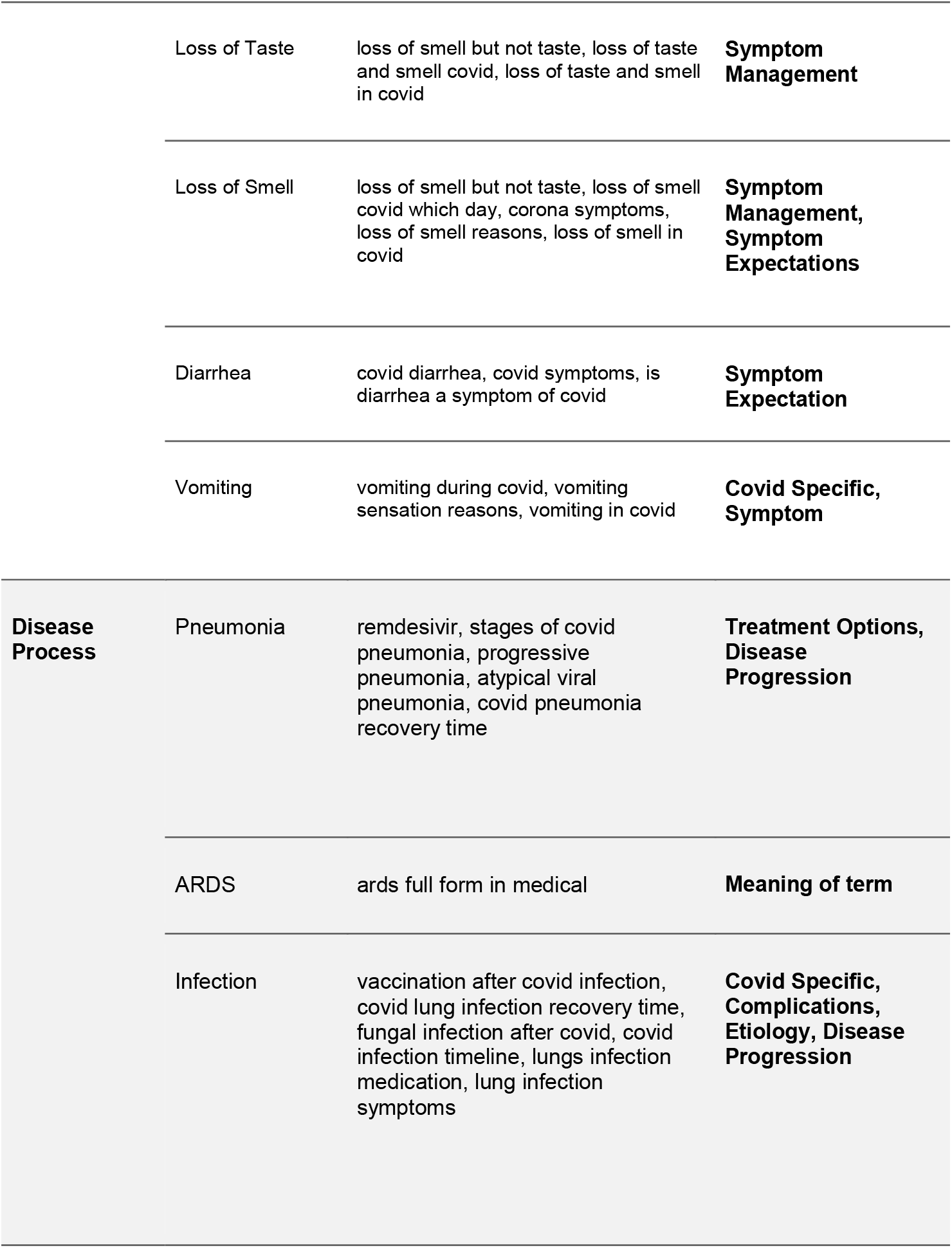
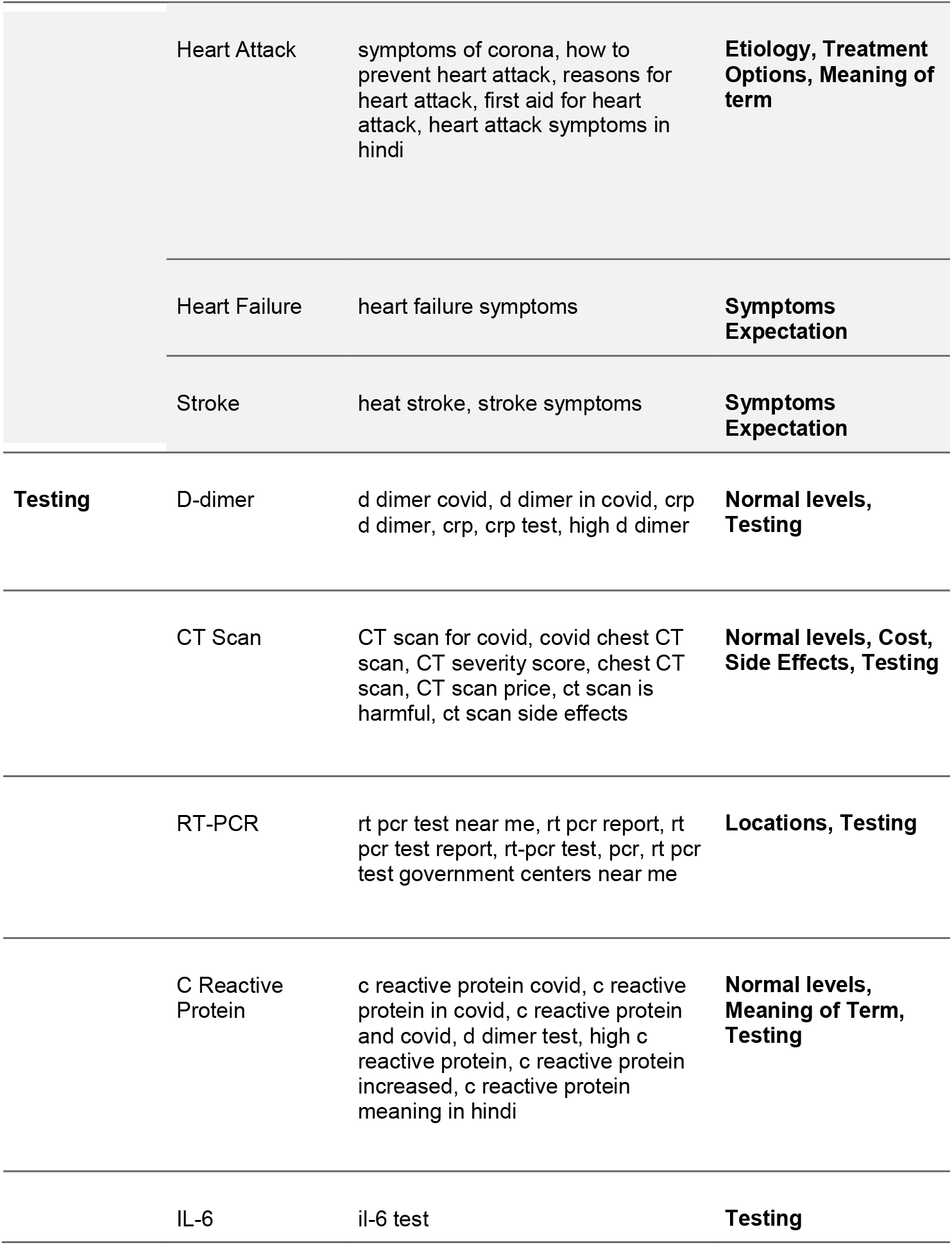
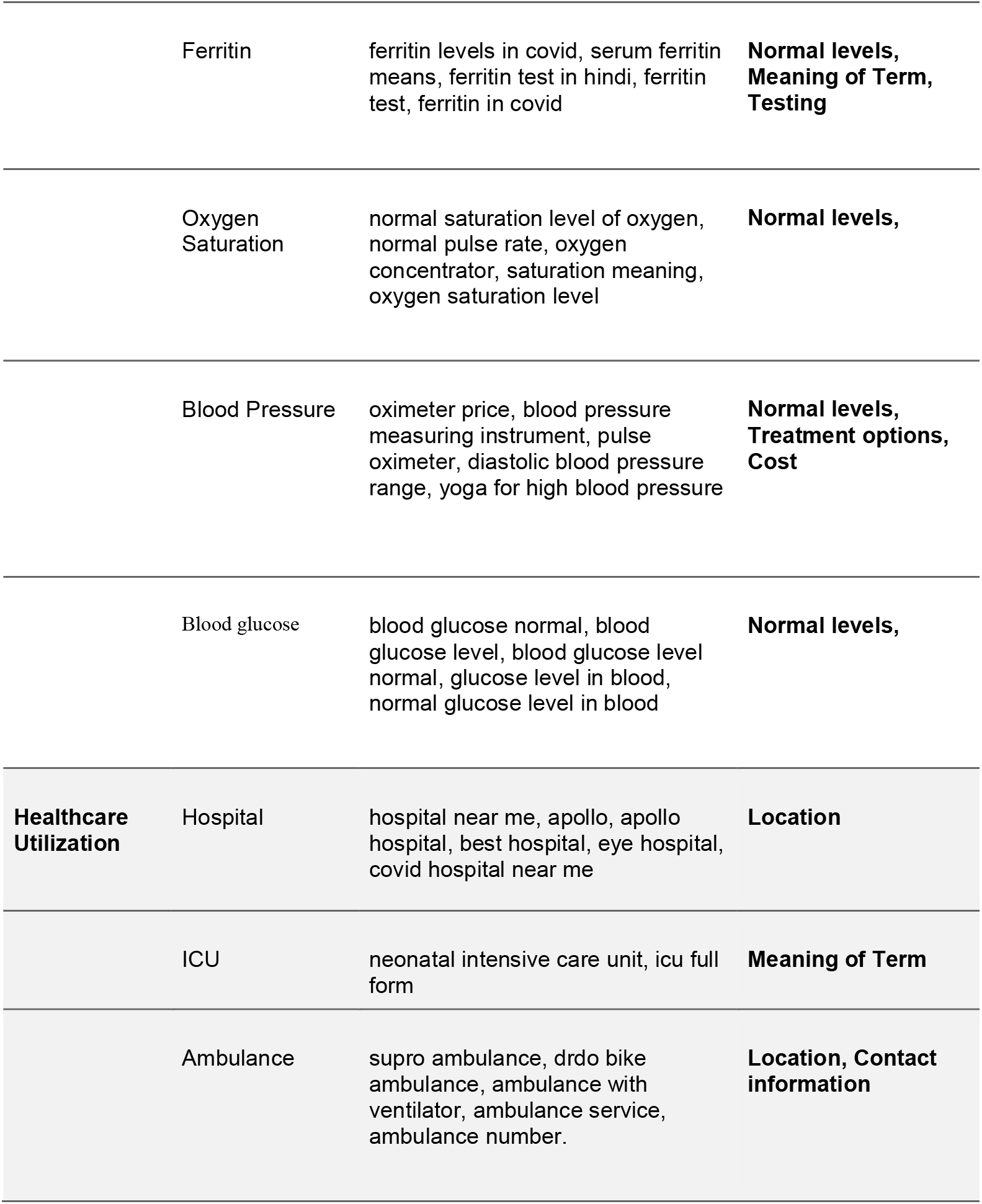
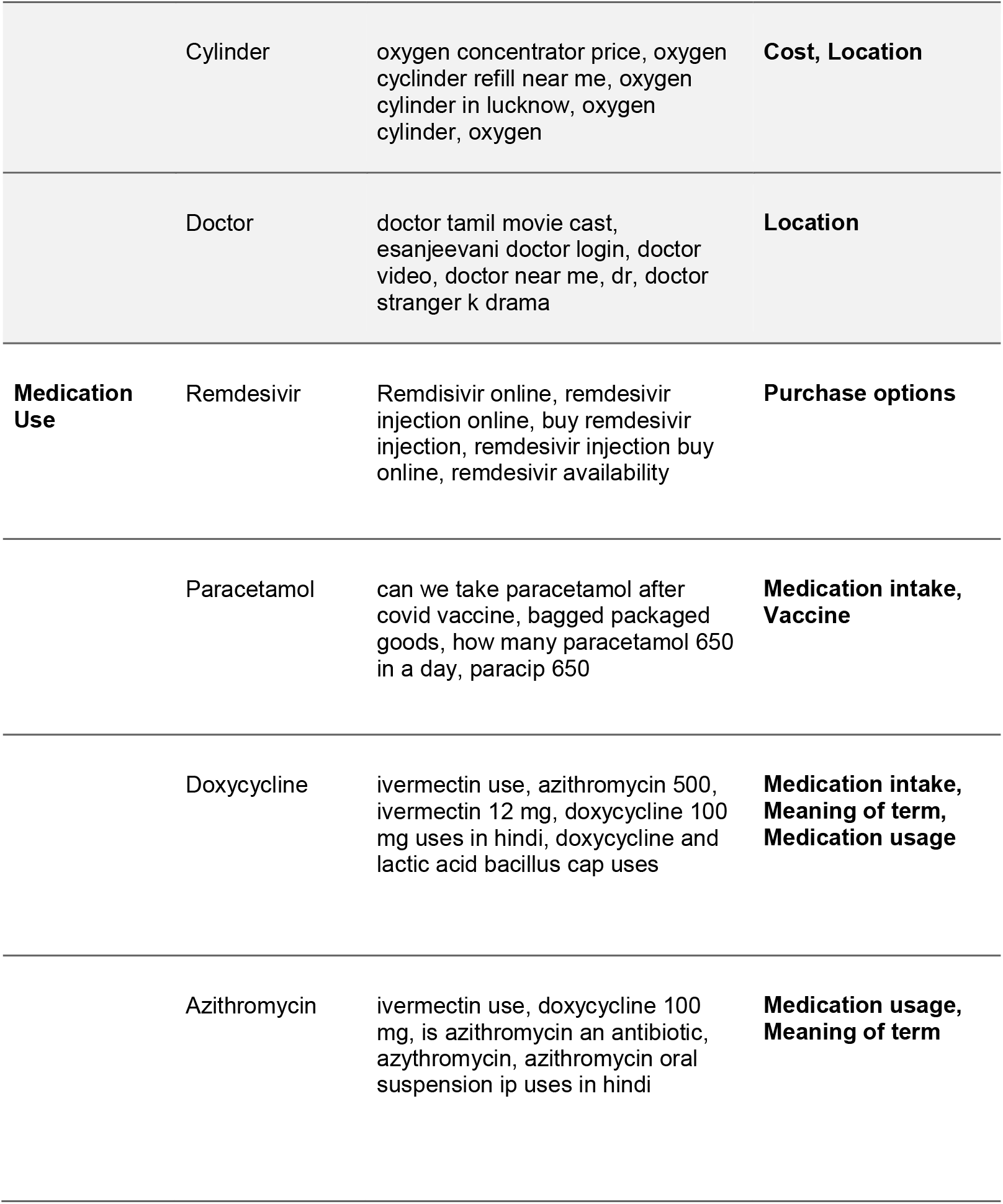
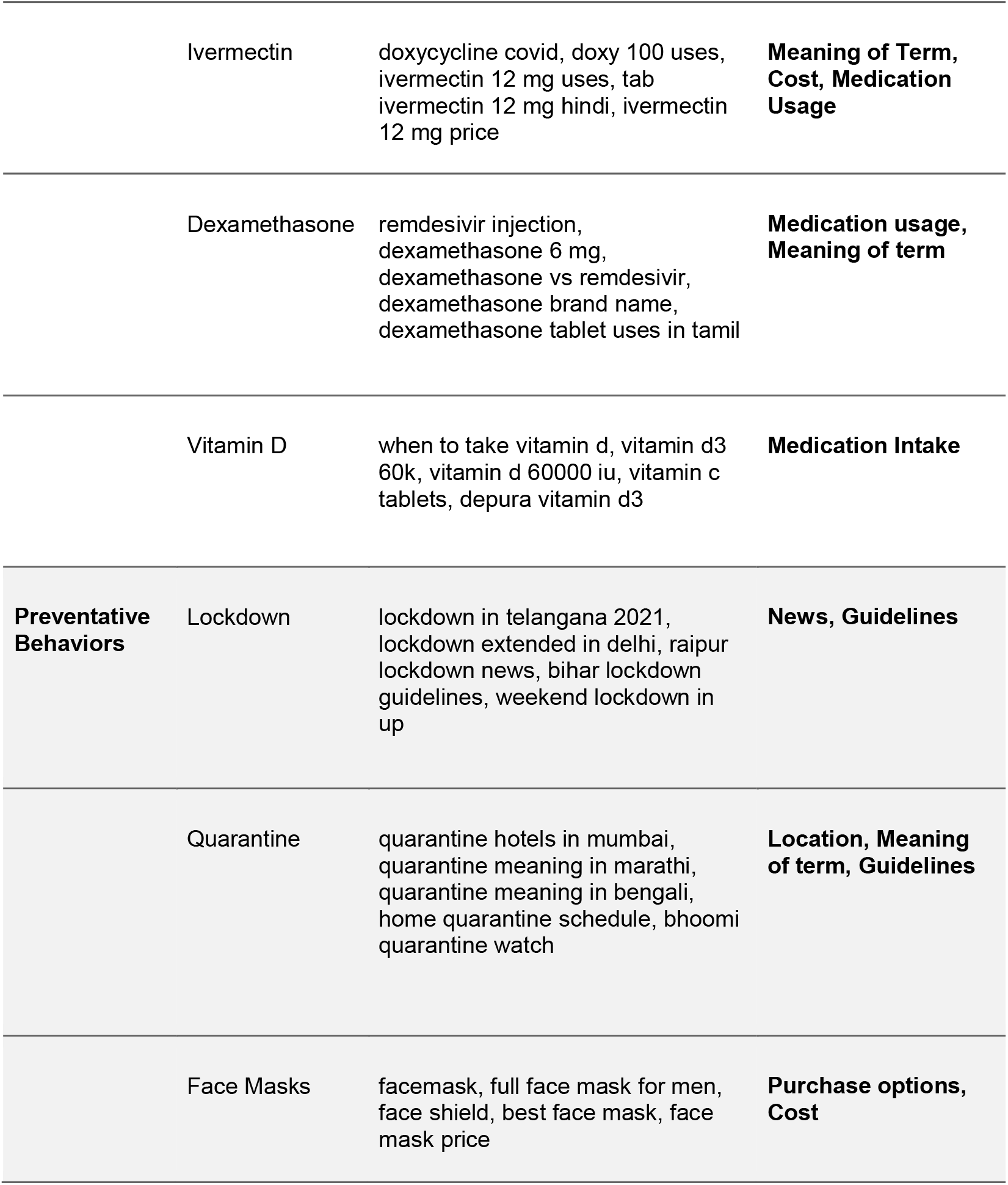
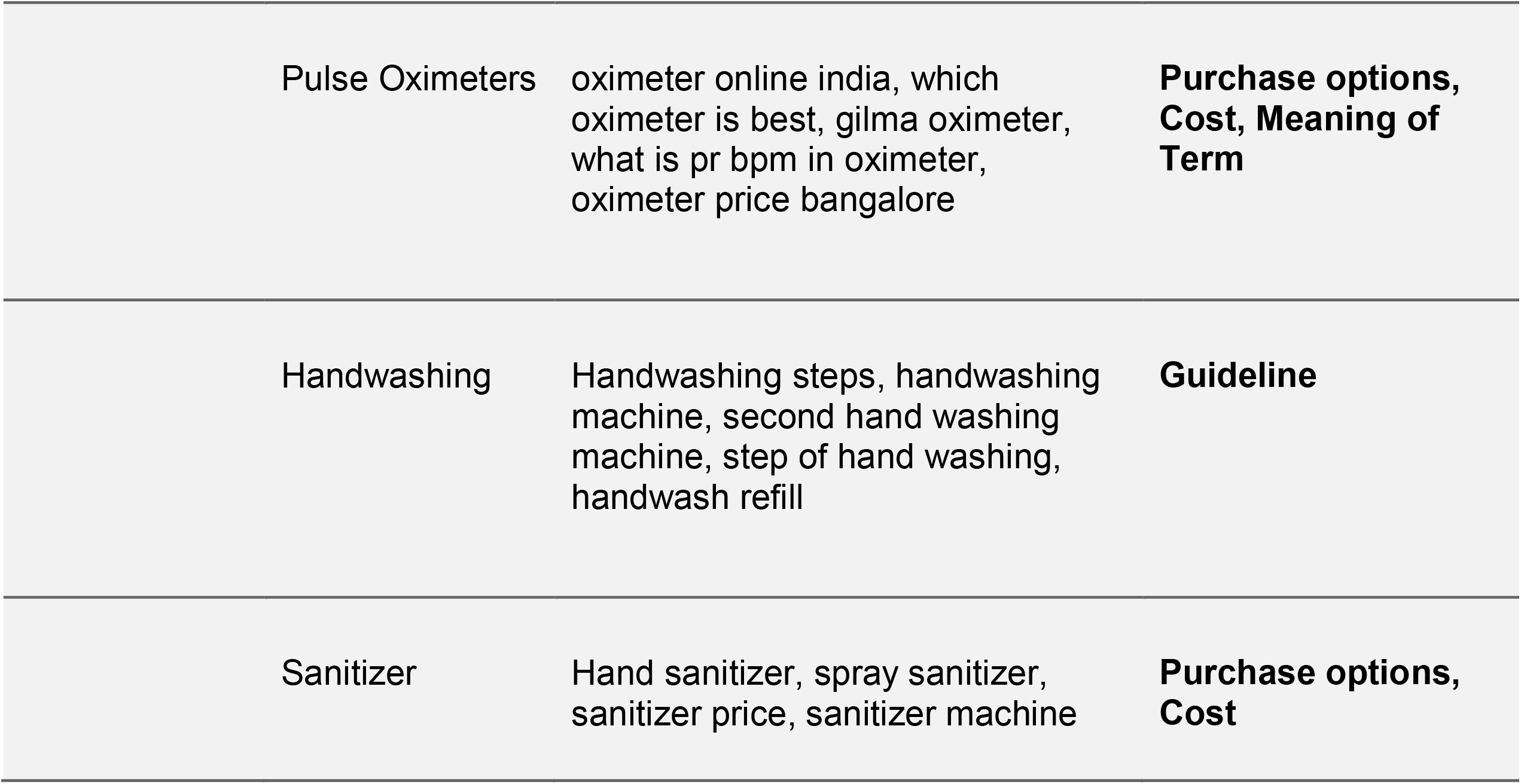
Thematic Analysis of Common Search Term-related Queries

## Discussion

In this retrospective, descriptive, ecological study of GT search terms related to the second wave of the COVID-19 pandemic in India, we observed that searches for symptoms (e.g. fever, cough), disease states (e.g. infection, pneumonia), COVID-19-related medications (e.g. remdesivir, ivermectin), testing modalities (e.g. PCR, CT Scan), healthcare utilization (e.g. oxygen cylinders, hospital), and preventive behaviors (e.g. pulse oximetry, lockdown) all demonstrated an increase over our time period from 2/12/21 - 5/9/2021, in line with increases in official case counts. While peak RSVs and doubling rates varied across search categories, we generally observed that symptoms, PCR testing, outpatient medications, and preventive behaviors peaked around April 24th, approximately two weeks prior to the peak RSV in official case counts, but exhibited a doubling date one-two weeks later than official case counts. Contrarily, healthcare utilization factors, including searches for hospital, physicians, beds, disease states, and inpatient medications did not peak until the first week of May. There was a highly significant correlation between ‘Coronavirus Disease 2019’, ‘Fever’, ‘Pulse oximetry’, ‘Oxygen saturation’, ‘C-reactive protein’, ‘D-Dimer’, & ‘Fabiflu’ and official case counts.

GT has been used to understand various aspects of the first wave of the COVID-19 pandemic worldwide, including its use as a digital surveillance tool [10-14], its use as a psychosocial behavior informant [15], and its use as a method to characterize novel symptoms such as ageusia and agnosia [16]. GT has many advantages, including real-time, real-world analysis, large sample size, and non-healthcare-related searches that may be missed in studies looking only at patients seeking healthcare.

We found that fever, cough, headache, fatigue, and chest pain were the most prevalent symptom-related searches during our time period, suggesting that these may be the most common presenting symptoms during the current COVID-19 wave. Searches for gastrointestinal symptoms, agnosia/ageusia, body/muscle aches, and other commonly reported symptoms from the first wave were not nearly as prevalent nor did they demonstrate a significant rise during our time period. These results are in line with current literature describing prevalence of symptoms [17]. Additionally, fever and cough demonstrated the strongest correlation with official case counts, whereas headache and fatigue showed significant, but lower, correlation and did not display a second doubling. This may suggest that either headache and fatigue are specific to certain patients, that there was already a baseline high level of RSV possibly due to ‘long-haul’ Covid [18], or that the pattern of symptom searching is biased away from headache and fatigue, which can be easily overlooked. The increase in searches for chest pain is likely the result of pleuritic chest pain from COVID-19 pneumonia rather than chest pain due to cardiovascular disease; however, Ciofani et al. showed in the U.S. population that hospital admissions for myocardial infarction decreased, while searches for chest pain increased in line with COVID-19 cases. This may suggest that patients may be self-triaging themselves or avoiding seeking care for true cardiovascular chest pain [19]. Interestingly, searches for fever and cough peaked nearly two weeks earlier than case counts, suggesting that symptomatic individuals may search symptoms prior to getting tested and being included in official case counts. If this is in fact true, GT may provide a useful tool to target populations with increases in searches for symptoms, by for example, providing testing facilities earlier in disease course.

These results demonstrate that GT may be useful in not only characterizing the symptoms of a novel population-wide disease but also in understanding the relative prevalence of certain symptoms among all individuals including those who otherwise have mild infection but may still search about fever or cough.

As severe COVID-19 infection is known to cause a pro-inflammatory state with elevated inflammatory markers and susceptibility to clot formation, we explored searches related to laboratory testing and disease complications including myocardial infarction, stroke, and pulmonary embolism. While searches for C-reactive protein and D-dimer were amongst the most highly correlated searches with official case counts - likely because PCR tests for disease are done in coordination with other laboratory tests and CT scans in suspected cases - we did not see a significant rise in myocardial infarction, stroke, or pulmonary embolism, suggesting that individuals with severe complications may not search before seeking care. Future studies should include terms for signs/symptoms of complications rather than the disease complication itself.

Our search term topics included commonly-used related terms (e.g. heart attack), but may not have been comprehensive enough to capture all related searches.

In addition to clinically-relevant searches, we also explored preventive behaviors and healthcare utilization. Searches for ‘oxygen saturation’, ‘pulse oximetry’, and ‘oxygen’ (cylinders) were highly prevalent and also significantly associated with official case counts. This is in agreement with the well-publicized issue of oxygen shortages in India and difficulties of finding hospital beds [20]. Despite the increase in searches for ‘hospitals’, we did not see a concomitant increase in searches for ‘ambulance’, making it unclear whether ambulance shortages were lower than hospital shortages, whether families were more likely to transport patients, or whether searches for hospitals were the result of media publicization rather than the primary cause.

In addition to GT’s use as a potential method to characterize disease, of equal importance is its role to better understand public questions concerning COVID-19 [21]. We found that related search queries could be grouped into three main themes, administrative, symptom expectations, and symptom management. This is in line with GT analyses by Hu et al. and Springer et al. that remarked that there is a need for increased disease awareness, with respect to treatment options and disease course [22,23]. This is especially evident and necessary in India, where health literacy remains low and many search queries were concerned with symptom management. There is also a need for India to address administrative questions including cost, location, and availability of testing, vaccine, and hospital facilities. Inability to access healthcare due to administrative issues is a primary source of disease burden and disproportionately affects those of lower socioeconomic status.

The results should be interpreted with caution due to many limitations. Firstly, our ecological study design does not allow us to make inferences at the individual patient level, nor does it provide information on the direction of correlation. Increases in searches may, at least partially, be due to increased presence of related topics in the media rather than individual situations. For example, Remdesivir exports were prohibited by Indian government officials on April 11th in anticipation of the predicted spike in demand for the drug. This may have contributed to the rise in interest for this term earlier than other medications.

Furthermore, searches for symptoms may not be solely due to COVID-19. However, given the high prevalence of COVID-19 In India and the significant correlation with confirmed COVID-19 cases, it is reasonable to infer that rises in related search terms are due to COVID-19. Additionally, search terms may not capture all non-English languages and regional colloquialisms. Although we tried to choose terms that would capture the largest percentage of related terms, this may have decreased specificity of searches. For example, the search topic “mask” rather than “face mask” or “covid mask” may encompass all of the intended searches with a high sensitivity; although the decision to choose the more general search term decreases specificity when including searches intended to find other masks, such as those for skincare or fashion purposes. Despite these limitations, GT’s large sample size, real-world and real-time benefits, and ability to capture populations that may otherwise be excluded from traditional research studies provide a useful tool to understand COVID-19. In particular, our major strength is there has been no comprehensive study on various clinical and social aspects of the second wave of the COVID-19 pandemic in India.

### Future Directions

Future studies should attempt to reconcile the predictive power of GT search terms, including highly correlated symptoms, testing, and healthcare utilization factors, in India, within specific regions in India, and other countries. Studies should also attempt to understand the impact of mass gatherings on GT searches to allow for prediction after upcoming events such as Eid. Future studies may also focus on understanding the association between search terms and official death counts in India, including prediction of underreporting of data based on GT models developed in other countries.

Furthermore, examination of other Internet activity besides Google Web Search (such as Youtube, Facebook, and Whatsapp) may provide a more accurate depiction of Internet-based health searches. In particular, Whatsapp is one of the largest modes of learning and spreading health-related information in South Asia [24]. Popular “chain-messages” or posts may be studied for keywords and interest based on time-course to check for correlation with COVID-19 case data and risk prediction capability.

## Data Availability

All data is available on trends.google.com. Our specific CSV file used for analysis can be provided upon reasonable request.

https://trends.google.com

